# Availability of Priority Maternal and Newborn Health Indicators: Cross-sectional Analysis of Pregnancy, Childbirth and Postnatal Care Registers from 21 Countries

**DOI:** 10.1101/2022.06.15.22276428

**Authors:** Mark Kabue, Francesca Palestra, Elizabeth Katwan, Allisyn C. Moran

## Abstract

**Background:** Data from national health management information systems (HMIS) are essential for routinely tracking progress toward national and subnational objectives, programmatic decision-making, planning and review, and ultimately to improve quality of care. It is through an in-depth understanding of the data elements captured in patient registers, as building blocks of national HMIS indicators, that it is possible to improve standardization in data collection and measurement of key indicators to help track progress towards achieving maternal and newborn health goals.

**Methods and findings:** This analysis was done through a review of antenatal (ANC), childbirth and postnatal (PNC) registers from 21 countries across five world regions. Between July and October 2020, various country-based maternal and newborn experts, implementing agencies, program managers, and ministries of health personnel were contacted and requested to share the current versions of the registers. Both paper-based and electronic registers were obtained. Twenty ANC registers, eighteen childbirth and thirteen PNC were analyzed. Both longitudinal and cross-sectional ANC and PNC registers were reviewed, while the childbirth registers included in the review were all cross-sectional. For the ANC registers 11/20 (55%) and for PNC registers 7/13 (54%) were longitudinal. In four countries the registers were electronic (Norway, Sweden, USA, West-Bank - Palestine), while the rest was paper-based. There was great variation in the number of data elements included from each register reviewed ranging from 24 in Nepal’s childbirth register to 188 in Eswatini’s ANC register. These findings are presented in four broad categories (Pregnancy, Childbirth, Postnatal care, and Mortality outcomes), with subheadings in each category.

**Conclusion:** This analysis highlights some areas in where there are data gaps as we work to reduce preventable maternal, newborn deaths and stillbirths and deliver high-quality, equitable care. Programme managers and health workers need this data to monitor the performance of their national health system and to guide the continuous improvement of health care services for women and newborns. The results of this analysis will complement additional work and help to inform the standardization of pregnancy and childbirth registers and provide information for other countries seeking to introduce indicators in their health systems.

## Introduction

The global burden of maternal deaths is estimated at 810 women dying each day from preventable conditions related to pregnancy and childbirth^1, 2^. Although there was 38% reduction in the maternal mortality ratio (MMR) between 2000 and 2017, the current MMR of 211 deaths per 100,000 live births remains unacceptably high especially, with two-thirds of the deaths occurring in low-to middle-income countries (LMICs)^1^. At the same time, the number of stillbirths is rising especially in Sub-Saharan Africa, increasing from 0.77 million in 2000 to 0.82 million in 2019. In some high-income countries – despite very low levels of neonatal mortality – more stillbirths than neonatal deaths occur, and in some cases, even surpass the number of infant deaths.^2, 3^. Most maternal and newborn deaths are preventable if proper preventive measures are in place. In many settings poor quality of care is a greater contributor to poor health outcomes than care coverage and it is estimated that about half of the maternal deaths and 58% of the newborn deaths could be averted with quality health care^4^. Maternal and newborn mortality is considered to be proxy indicator of limited access to quality healthcare service delivery in a population. Avoidable maternal and newborn health inequalities arise because of the socio-cultural and socio-eco-political circumstances in which a woman and her newborn grow, live, work, and age, and the systems put in place to deal with illness^5^.

The data needed for routine program implementation monitoring and evaluation are gathered through a variety of modalities, with health facility registers being an important source of information to document events around antenatal care (ANC), childbirth, and postnatal care (PNC), hence the need to periodically assess their content and utility^6^. However, there are challenges related to using the current disjointed data collection and reporting systems which in part included focus on meeting the needs of specific funders, poorly developed health information systems (HIS), and inadequate human resources capacity^7^. Furthermore, health data are often recorderd and/or collated by clinicians who have to deal with high patient caseloads especially those working in the public sector, which might lead to staff burnout, compromised data quality and well-being of the patients^8^. Health facility data have been reported to have many shortcomings which include poor quality, inaccuracies, incompleteness and not reporting on time through the routine health information systems^9–16^.

National health management information systems (HMIS) are also essential for decision-making aiming to improve the quality of care, and routinely tracking progress toward national and subnational objectives, including patient management objectives, for which data cannot be collected otherwise^17^. The majority of national maternal and newborn health indicators can be reported through the HIS (and are used by countries to track their progress toward local and national goals). One such commonly used indicator which serves as a marker of service utilization is proportion of women attending at least four ANC visits during pregnancy. Data elements used to compute this indicator are captured at the facility level and reported monthly by all facilities using the established reporting channels in the country, often flowing from facility to subnational and then to national level, with consolidation of data along at the national level^19, 20^.

RHIS is often weak in most low- and middle-income countries, with limitations in the representativeness and quality of routine and health facility data^17^. In many LMICs countries, routine data are captured through use of printed paper registers, and only available through electronic platforms in summary form, using software such as the district health information system (DHIS2)^21, 22^. On the other hand, high-income countries such as Denmark, Norway, Sweden, and United States of America have robust electronic data capture systems^23–26^.

Analysis of antenatal care, childbirth and postnatal care registers is an important step towards gaining a deeper understanding of key data elements related to maternal and newborn health in the national HMIS across the world with a view towards promoting standardization in data collection, analysis, and use^27^. This paper reports on analysis of key data elements in registers of countries whose data sources were collected and reviewed, describes differenes in the way data elements are captured, and highlights opportunities for inclusion of some critical data elements that are not currently collected in registers.

### Methodology

The approach used in this review of registers was informed by previous similar work that focused on maternal, newborn and child survival^13, 27^. Selection of the countries to be included in the review was guided by the findings of literature review conducted prior to embarking on the analysis of the registers as well as the availability of the records. Countries included in this review had some literature published related to antenatal care, childbirth, or postnatal care registers. A few countries especially those from regions where published literature was not available, were added to the list to present a more global perspective, taking into account the geographical distribution. In the end, twenty-one countries drawn from five regions (Africa, Europe, South-East Asia, Eastern Mediterranean, and the Americas), were included in the review (Table 1).

**Table 1:** Countries whose registers were included in the review.

| Country |  | WHO Region | Version-year of registers | Format: Paper or Electronic | Antenatal care registers | Postnatal care registers |
| --- | --- | --- | --- | --- | --- | --- |
| 1 | Afghanistan | Eastern Mediterranean | 2018 | Paper | L* | L |
| 2 | Argentina | Americas | 2018 | Paper | L | X** |
| 3 | Bolivia | Americas | 2018 | Paper | X |  |
| 4 | Burkina Faso | Africa | Unspecified | Paper |  | X |
| 5 | Eswatini | Africa | 2015/6 | Paper | L | L |
| 6 | Ghana | Africa | 2015 | Paper | X | L |
| 7 | Guyana | Americas | 2015 | Paper | L |  |
| 8 | Indonesia | South-East Asia | 2019 | Paper | L | L |
| 9 | Kenya | Africa | 2019 | Paper | X | X |
| 10 | Malawi | Africa | 2018 | Paper | L | L |
| 11 | Nepal | South-East Asia | Unspecified | Paper | X |  |
| 12 | Nigeria | Africa | 2019 | Paper | X | X |
| 13 | Norway | Europe | 2018 | Electronic | L | L |
| 14 | South Sudan | Africa | 2015 | Paper | X | X |
| 15 | Sweden | Europe | 2016 | Electronic | L |  |
| 16 | Tanzania | Africa | 2019 | Paper | X | X |
| 17 | Uganda | Africa | 2019 | Paper | X |  |
| 18 | USA | Americas | N/A | Electronic | L |  |
| 19 | West Bank - Palestine | Eastern Mediterranean | N/A | Electronic (Pilot) | L |  |
| 20 | Zimbabwe | Africa | 2019 | Paper | X |  |
| 21 | Zambia | Africa | 2020 | Paper | L | L |

\*L denotes longitudinal register; \*\*X denotes cross-sectional register. Grey areas indicate that the registers were not available for analysis.

#### Gathering the Registers

Between July and October 2020, various country-based maternal and newborn experts, implementing agencies, program managers, and ministries of health personnel were contacted and requested to share the current versions of the ANC, childbirth, and PNC registers available to them. These materials were shared in various formats such Microsoft Excel and Word, portable document format (PDF), images, and links to websites. Both paper-based and electronic registers were obtained from 21 countries through different methods, primarily electronically. For the paper-based registers, the years of their production ranged from between 2015 to 2020. For some of the electronic registries, no specific dates could be assigned to when they were produced. However, majority had some time-periods documented in various ways; for example, the medical birth register of Sweden reviewed spans the period 1967 to 2018^23, 24, 28^.

For materials that were not in English, these were translated into English prior to review and data abstraction. Since the registers are revised periodically, the most recent versions of the documents available for each country were analyzed. The ANC and PNC registers reviewed were in both cross-sectional and longitudinal (i.e. All visits for an individual recorded on the same page in the register) formats. 11/20 (55%) of ANC registers and 7/13 (54%) of PNC registers analyzed were longitudinal. All childbirth registers reviewed were cross-sectional (table 1). Registers even from the same country were not linked to each other; they were siloed by the type of care.

#### Capturing the Data Elements

A Microsoft Excel worksheet was used to store key data elements manually extracted from each register foreach country. These data included; name of the data source, year of production, format (cross-sectional or longitudinal), and the data elements contained. For each country file, separate worksheets were generated for ANC, childbirth (or labour and delivery), and PNC data elements. Thereafter, all data elements contained in the register were marked under the appropriate worksheets, organized in logical sections such as demographics, medical history, laboratory investigations, client outcomes and/or services offered. Abstraction of the information was done in a standardized manner by one person (MK) over a period of three months. Data quality checks were conducted through a second review of select registers from the 21 countries.

#### Analysis and Synthesis of Content

Following completion of the data abstraction into the individual country worksheet, a summary file with three worksheets (ANC, childbirth, PNC) was generated. This process was done in a stepwise manner as follows:

**Step 1** – Five categories were generated for summarizing the data, and corresponding worksheets in the Microsoft Excel file generated: Pregnancy, Lab tests and investigations, Birth outcomes and treatment, and PNC.

**Step 2** - Country names were listed alphabetically, forming the rows.

**Step 3** - The column headings were generated by identifying the common data elements present in a majority of the registers. Some key data elements that are either denominators or numerators of key maternal and newborn health indicators were also included, whether they were present in many registers or not..

**Step 4** – Summary data were captured from each country file by marking a “check” in the appropriate cell if the data element or term listed on the column heading was present in the country file.

**Step 5** – The content of the data elements captured was further categorized into logical categories, guided by standard definitions of key maternal and newborn indicators.

**Step 6** – As the process of capturing the summary data progressed, some data elements were added and steps 3-5 were reviewed, with corresponding adjustments being made to the data summary categories identified.

**Step 7** – Sub-analysis of registers from four countries (Ghana, Kenya, Nigeria, and Zambia) across two time periods (2017/8 and 2019/20) was done to identify changes in format and content or data elements included. These countries were selected because data on their registers were available from prior similar review conducted in 2018^11^; this enabled comparison of data between two points in time. We looked for the presence or absence of twenty-seven data elements that are critical to monitoring maternal and newborn services across the continuum of care, from pregnancy to PNC.

## Results

The findings on the analysis of the similarities and differences on how key data elements are captured in different registers from 21 countries are presented. There was great variation in the number of data elements included from each document reviewed ranging from 24 in Nepal’s delivery register to 188 in Eswatini’s ANC register. These findings are presented in three broad categories (Pregnancy, Childbirth, Postnatal care, and Mortality outcomes), with subheadings in each category.

### Pregnancy

Of the 20 countries whose ANC registers were reviewed, age, name of the woman, ANC number assigned at the facility or a personal identifier, and residential address were the most commonly recorded data elements (Figure 1). In a few countries, information on the husband or baby’s father including name (Afghanistan) and citizenship (Sweden) is recorded. Some other vital information recorded, usually during the first ANC visit, include woman’s relevant medical history such as hypertension and diabetes in 13/20 (65%) of the countries, and complications from past pregnancies in 7/20 (35%) of the countries. Recording of behavioural or practices such as smoking, use of alcohol or drugs is documented in in five countries, all located either in Europe or the Americas.

**Figure 1:**
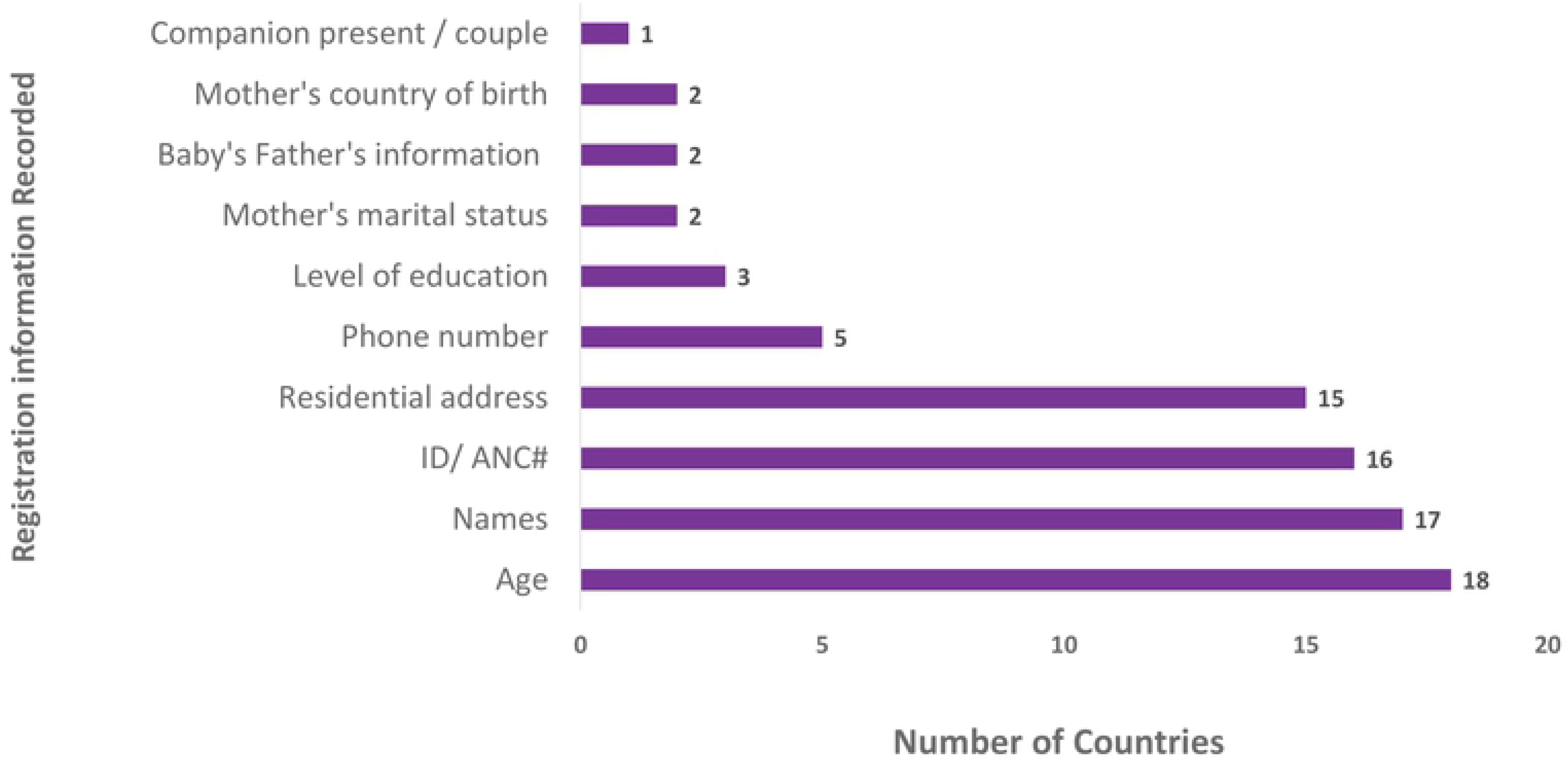
Client registration and baby’s father’s information.

#### Utilization of antenatal care services

Countries that used longitudinal ANC registers routinely document between five and eight or more visits, whereas those using electronic formats (Norway, Sweden, USA, and West Bank – Palestine), have capacity to record many more visits, one visit per row. For the countries using cross-sectional registers, they record the first visit as “New visit” and subsequent visits in a separate column as “Revisit”. In some cases, the actual visit number was entered, for example 3^rd^ visit, while in some cases, there was a column to capture 4+ visits in 8/20 countries.

#### Routine examination during ANC visits

During each ANC visit, a variety of measurements are taken and recorded, allowing for comparison of a current visit to previous visits in order to identify or monitor possible early danger signs. In over half of the countries examined, routine data elements recorded during a given visit included: gestational age, blood pressure, weight, height, and haemoglobin level or pallor/ anaemia. The less commonly recorded parameters included: fetal heart tones/sounds, presentation of the baby, and the mother’s nutrition status (Table 2).

**Table 2:**
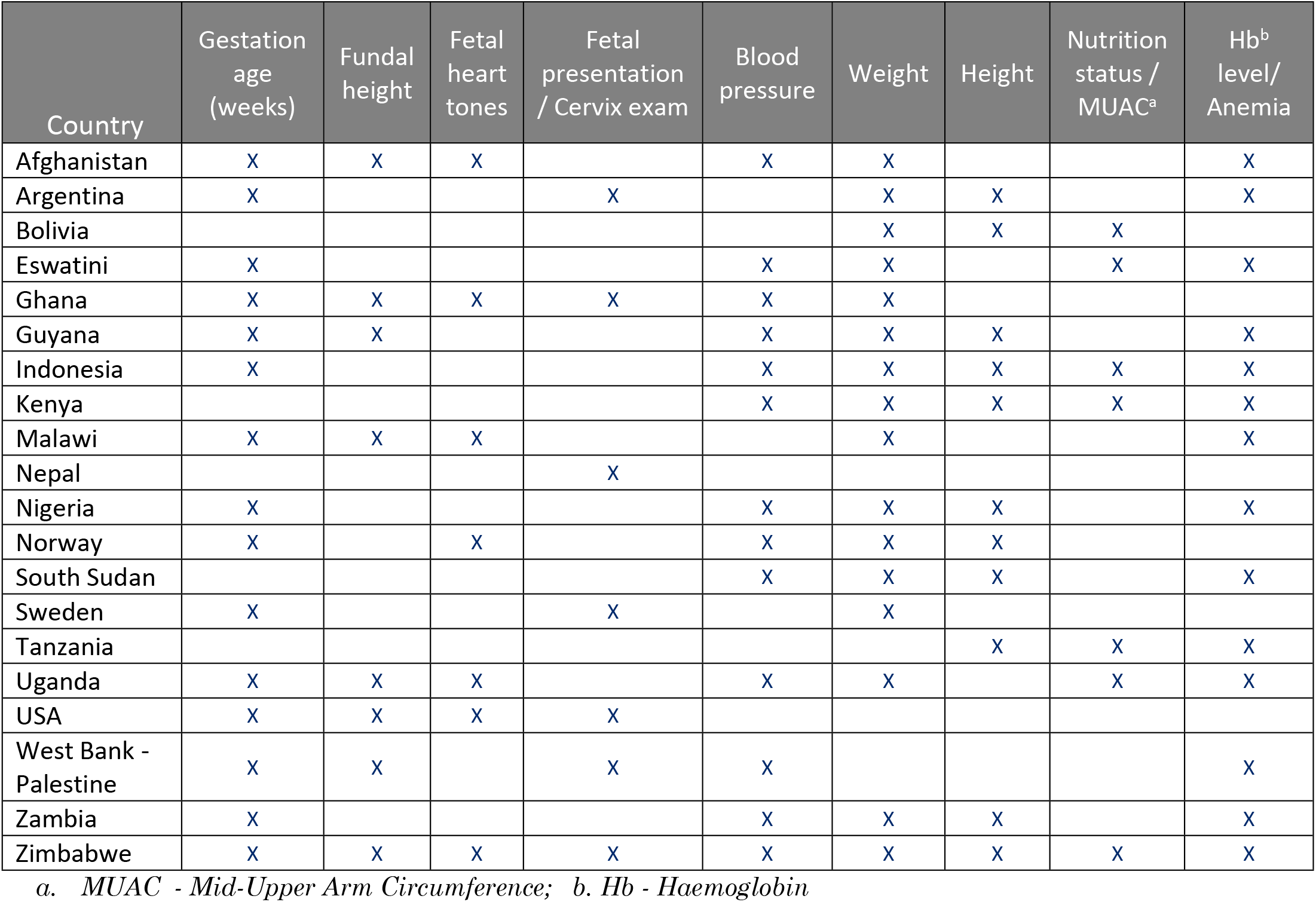
Key measurements during antenatal care visits.

| Country | Gestation age (weeks) | Fundal height | Fetal heart tones | Fetal presentation / Cervix exam | Blood pressure | Weight | Height | Nutrition status / MUAC<sup>a</sup> | Hb<sup>b</sup> level/ Anemia |
| --- | --- | --- | --- | --- | --- | --- | --- | --- | --- |
| Afghanistan | X | X | X |  | X | X |  |  | X |
| Argentina | X |  |  | X |  | X | X |  | X |
| Bolivia |  |  |  |  |  | X | X | X |  |
| Eswatini | X |  |  |  | X | X |  | X | X |
| Ghana | X | X | X | X | X | X |  |  |  |
| Guyana | X | X |  |  | X | X | X |  | X |
| Indonesia | X |  |  |  | X | X | X | X | X |
| Kenya |  |  |  |  | X | X | X | X | X |
| Malawi | X | X | X |  |  | X |  |  | X |
| Nepal |  |  |  | X |  |  |  |  |  |
| Nigeria | X |  |  |  | X | X | X |  | X |
| Norway | X |  | X |  | X | X | X |  |  |
| South Sudan |  |  |  |  | X | X | X |  | X |
| Sweden | X |  |  | X |  | X |  |  |  |
| Tanzania |  |  |  |  |  |  | X | X | X |
| Uganda | X | X | X |  | X | X |  | X | X |
| USA | X | X | X | X |  |  |  |  |  |
| West Bank - Palestine | X | X |  | X | X |  |  |  | X |
| Zambia | X |  |  |  | X | X | X |  | X |
| Zimbabwe | X | X | X | X | X | X | X | X | X |

*a. MUAC - Mid-Upper Arm Circumference; b. Hb - Haemoglobin*

#### Malaria in pregnancy

In countries where malaria is endemic, documentation of the doses of intermittent preventive treatment in pregnancy (IPTP) for management of malaria in pregnancy was included in 7/20 (35%) of the registers reviewed. Four of the seven countries document at least four doses while the other three record three doses. Provision of bed nets was included in 9/20 (45%) of the ANC registers reviewed (Table 3). Two countries had columns to document malaria treatment provided.

**Table 3:** IPTp doses, Malaria Treatment and Bednets.

| Country | IPTp1 | IPTp2 | IPTp3 | IPTp 4 or more | Malaria treatment | LLIN/ ITN provided |
| --- | --- | --- | --- | --- | --- | --- |
| Afghanistan | X | X | X | X | X | X |
| Ghana | X | X | X |  |  | X |
| Indonesia |  |  |  |  | X | X |
| Kenya | X | X | X |  |  | X |
| Malawi | X | X | X |  |  | X |
| Nigeria | X | X | X | X |  | X |
| South Sudan |  |  |  |  |  | X |
| Tanzania | X | X | X | X |  |  |
| Uganda |  |  |  |  |  | X |
| Zambia | X | X | X | X |  | X |

#### Prevention of mother to child transmission (PMTCT) of HIV and STIs

Pregnancy is a period when specific measures can be taken to prevent a baby born to a HIV-infected woman from being infected. In addition, screening and treating mothers with sexually transmitted diseases like syphilis can protect the baby from such infections both in-utero or after birth. The HIV status of the mother was documented in 10/20 (50%) of countries where ANC registers were reviewed. ANC registers from nine countries (45%) document whether the mother is on antiretroviral therapy (ART). Three countries (Eswatini, Kenya, and Uganda) document the WHO stage of disease. The same three countries also have space to document whether the pregnant woman is on cotrimoxazole to prevent opportunistic infections. Regarding syphilis, 7/20 (35%) of the countries record treatment of syphilis (Table 4).

**Table 4:** Screening and testing performed during antenatal care visits.

| Country* | Urinalysis | Hb test | Blood sugar | Blood group | Rh factor | Syphilis Testing | HIV Testing | Hep. B Testing | CD4 Count: | VL | Partner HIV Testing: | HIV Couple Testing: | Malaria testing | TB test | Pap Smear | Ultrasound |
| --- | --- | --- | --- | --- | --- | --- | --- | --- | --- | --- | --- | --- | --- | --- | --- | --- |
| Afghanistan | X | X | X | X | X | X |  |  |  |  |  |  |  | X |  |  |
| Argentina | X | X | X | X | X |  | X |  |  |  |  |  |  |  |  |  |
| Bolivia |  |  |  |  |  |  |  |  |  |  |  |  |  |  | X |  |
| Eswatini | X | X |  | X | X | X | X |  | X | X | X | X |  |  |  |  |
| Ghana |  |  |  | X | X | X |  | X |  |  |  |  |  |  |  |  |
| Guyana | X | X |  | X | X | X | X | X |  |  |  |  |  |  | X | X |
| Indonesia | X | X | X |  |  | X | X |  |  |  |  |  | X | X |  |  |
| Kenya | X | X |  | X | X | X | X |  | X | X | X | X |  | X |  |  |
| Malawi | X | X |  |  |  | X | X |  |  |  |  |  |  |  |  |  |
| Nepal |  |  |  |  |  | X | X |  |  |  | X |  |  |  |  |  |
| Nigeria | X | X | X |  |  | X | X | X |  |  |  |  |  |  |  |  |
| Norway* |  |  |  |  |  |  |  |  |  |  |  |  |  |  |  |  |
| South Sudan | X | X |  | X | X | X | X |  |  |  | X | X |  |  |  | X |
| Sweden |  |  |  |  |  |  |  |  |  |  |  |  |  |  |  | X |
| Tanzania | X | X | X |  |  | X | X |  |  |  |  |  | X |  |  |  |
| Uganda | X | X |  | X | X | X | X |  | X | X | X |  |  |  |  | X |
| United States |  | X |  | X | X | X | X |  |  |  |  |  |  | X | X |  |
| West Bank - Palestine | X | X | X | X | X |  |  |  |  |  |  |  |  |  |  | X |
| Zambia | X | X | X |  |  | X | X | X |  |  | X | X |  | X |  |  |
| Zimbabwe | X | X |  | X | X | X | X |  | X | X | X |  |  | X |  | X |

\* Norway's electronic pregnancy register did not have data on laboratory tests performed during pregnancy; probably these data are captured through a different system.

#### Nutrition and micronutrients supplementation

Nutrition interventions are key to ensuring the good health of the mother and normal growth of the unborn baby. Provision of health promotion and prevention messages to the mother through nutrition counseling is documented in 9/20 (45%) of the countries. Regarding micronutrients, six countries document provision of iron and folate supplementation separately (Guyana, Indonesia, Kenya, Norway, Tanzania, Uganda, and Zambia), while eight countries have them combined in one column. Provision of Vitamin A and calcium are recorded in one country each, Norway and Zimbabwe, respectively. Related to nutrition, deworming is recorded in only six countries (Figure 2).

**Figure 2:**
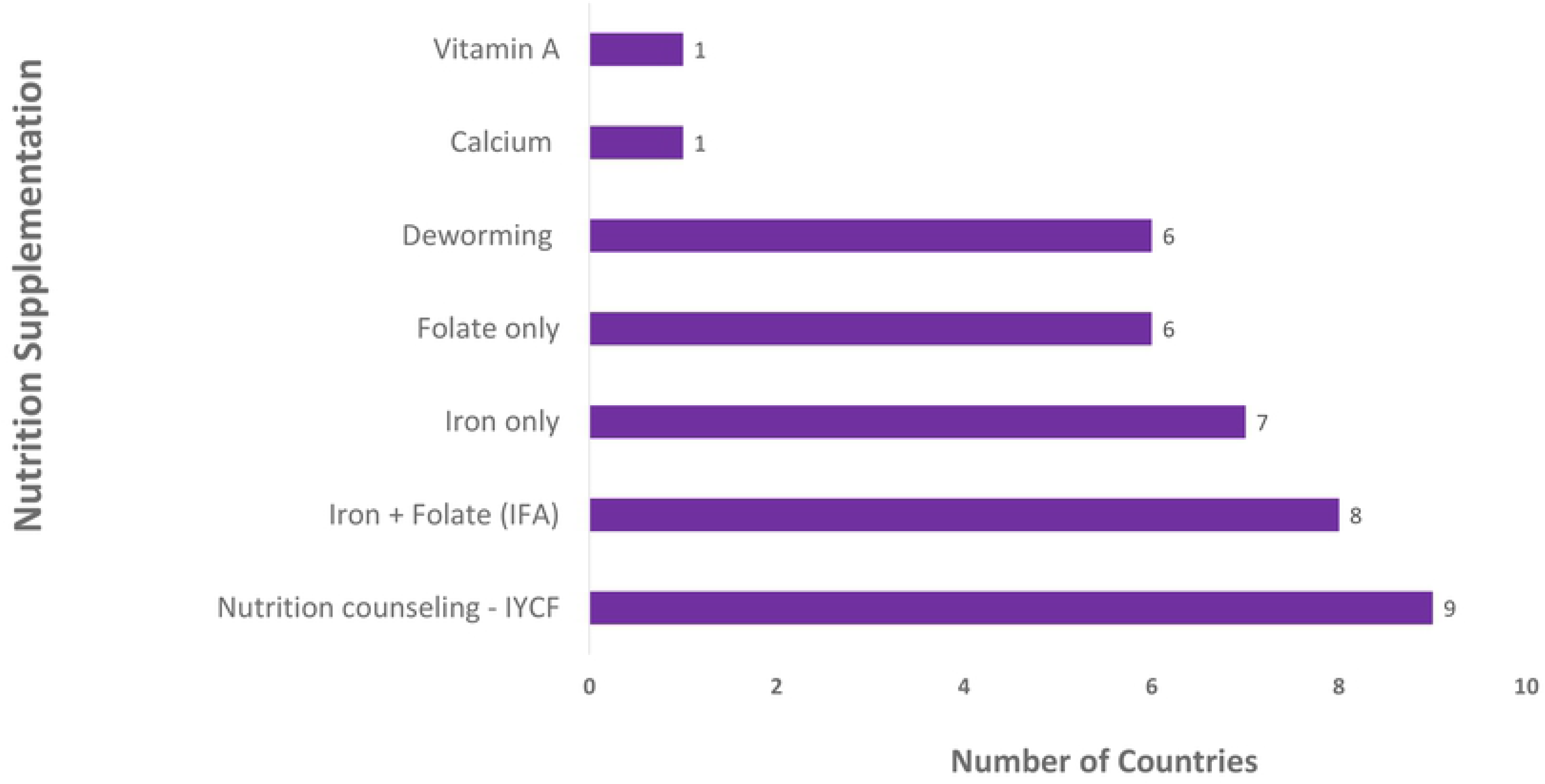
Nutrition supplementation and counseling.

#### Family planning counselling and screening

Family planning counseling provided during pregnancy was documented in five countries. At the same time, discussion around the birth plan is recorded in five countries (Afghanistan, Argentina, Sweden, USA, and Zimbabwe). Screening for gender-based violence (GBV) is documented in Afghanistan, while breast cancer screening though physical examination is recorded in Zambia.

#### Routine Laboratory tests and other investigations done during ANC

Majority of the countries capture data on laboratory tests and investigations conducted routinely most of them during each ANC visit. The most commonly recorded tests in the countries reviewed are: hemoglobin level; Syphilis testing; Urinalysis for sugar or proteins; HIV test; and blood grouping as shown in table 4. Six countries document Tuberculosis screening. Hepatitis B testing is recorded in four countries. Pap smear is one of the investigations done that are documented only three countries. Ultrasound and status of the baby is recorded in six countries.

### Childbirth

The care received by the mother from the moment of admission to discharge after delivery is recorded mostly in a maternity register. However, some other information such as the personal identifying information of the mother medical history is recorded in supplemental documents such as ANC card or other records retained by the client. The estimated date of delivery and last known menstrual period dates are not consistently documented in the maternity register.

#### Monitoring of mother and the fetus during labor

Eighteen country childbirth registers or source documents were reviewed. The information captured in the registers distinguished between vaginal deliveries, assisted deliveries, and cesarean section deliveries in the register. Information in the register on whether a partograph was used is recorded in four countries, as shown in table 5.

**Table 5:** Childbirth information.

| Country | Onset of delivery | Date of Delivery | Time of Delivery | Duration of labour | Use of Uterotonic | Pantograph used | Mode of Delivery |
| --- | --- | --- | --- | --- | --- | --- | --- |
| Afghanistan | X | X | X |  | X |  | X |
| Argentina |  |  |  |  | X |  |  |
| Bolivia |  |  |  |  |  |  | X |
| Eswatini |  | X | X |  |  |  | X |
| Ghana |  |  |  |  | X |  | X |
| Guyana* |  |  |  |  |  |  |  |
| Indonesia | X | X | X |  | X | X | X |
| Kenya |  | X | X | X | X |  | X |
| Malawi | X |  |  |  | X |  |  |
| Nepal | X |  |  |  |  |  |  |
| Nigeria |  | X | X |  | X | X | X |
| Norway | X | X | X | X |  |  | X |
| South Sudan | X |  |  |  | X |  |  |
| Sweden | X | X | X |  |  | X | X |
| Tanzania | X | X | X | X | X |  | X |
| Uganda |  | X |  |  | X |  | X |
| West Bank - Palestine |  |  |  |  |  |  | X |
| Zambia | X | X | X | X | X | X | X |

\* Guyana register did not capture data on any of the six key data elements related to childbirth, but had space to capture data on name of the service provider and other treatments. Burkina Faso and Zimbabwe did not have childbirth registers to be analyzed.

The data elements in the partograph were not part of this review. Use of a uterotonics (Oxytocin, Ergometrine or Misoprostol) immediately after delivery is documented in 11/18 (61%) countries, while use of magnesium sulphate in the management of pre-eclampsia/ Eclampsia is recorded in three countries. Complications of Childbirth are documented in 14/18 (78%) countries. The complications documented include vaginal tears, blood loss or need for transfusion, retained placenta, obstructed, or prolonged labor, ruptured uterus, and sepsis (Figure 3).

**Figure 3:**
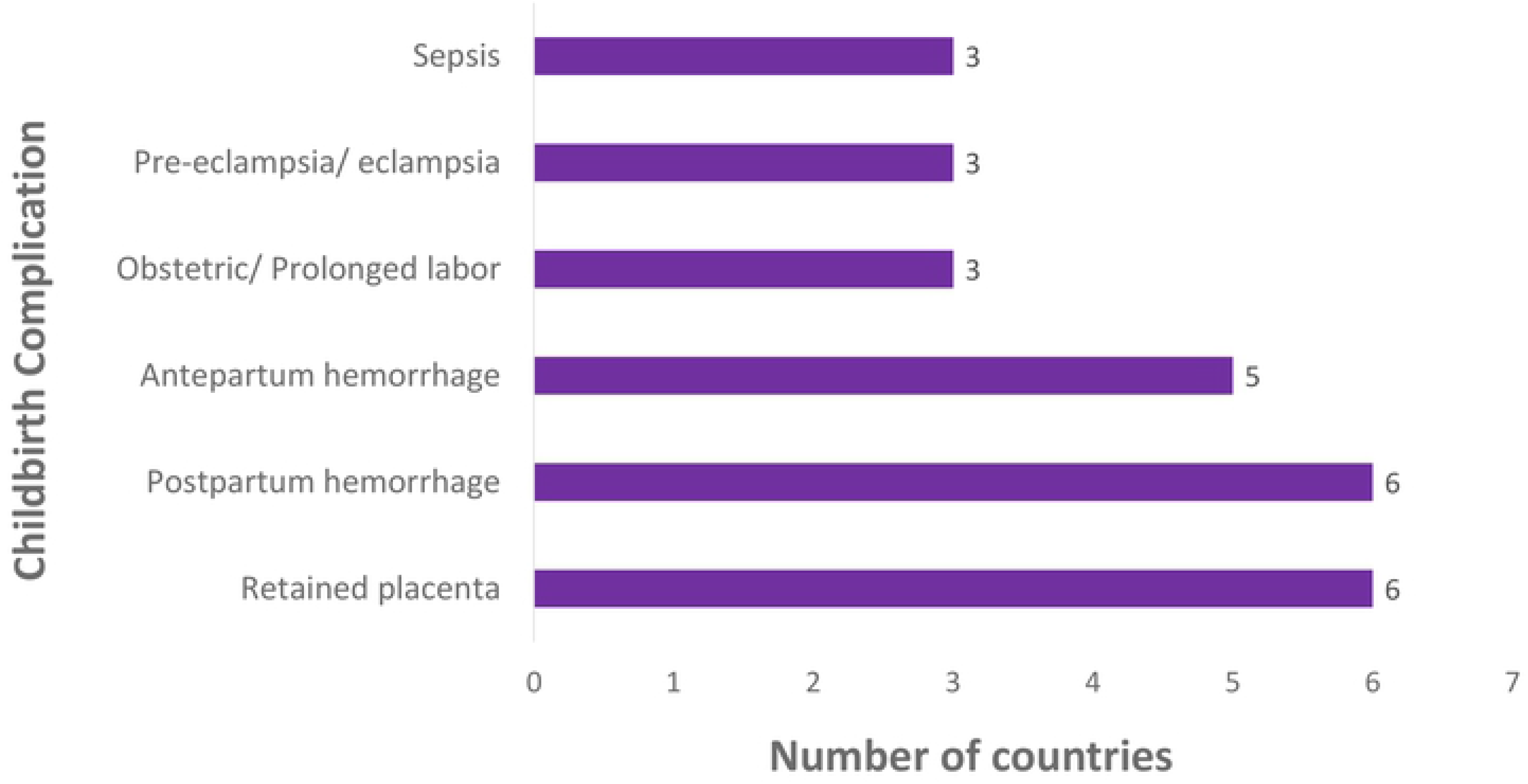
Maternal complications of childbirth.

#### PMTCT during childbirth

The labor and delivery period is another window of opportunity to implement PMTCT, similar to the pregnancy period. However, documentation of HIV status of the mother is not commonly included in many maternity or childbirth registers. In this review, five countries (Afghanistan, Eswatini, Guyana, Kenya, and Malawi) capture data on either HIV testing at delivery or the HIV status at admission. Partner HIV status is even less documented, only in Kenya and Malawi. Similarly, very few countries document HIV testing for the baby, CD4 count and viral load monitoring of the mother in the childbirth registers (only Kenya and Uganda). Guyana and Kenya are the only countries that record syphilis testing in childbirth.

#### Best practices in newborn care

The newborn’s first hour of life requires key lifesaving interventions to implemented immediately postpartum to ensure the survival of the baby. For newborns experiencing difficulty in breathing at birth, resuscitation is documented in eight countries while breastfeeding within the first hour is recorded in six countries out of the 18 childbirth registers reviewed. The practice of skin-to-skin is documented in Kenya, and immediate cord care through application of either chlorohexidine or chloramphenicol is recorded in Ghana, Kenya, and Nigeria.

In 15 out of the 18 countries, live births are clearly recorded, distinguishing them from fetal and newborn death. The sex and weight of the baby are recorded in 10 and 12 countries, respectively. The Apgar score is recorded in 10 countries, with most of them categorizing by the time the assessment was done; one, five, or 10 minutes after birth. Recording of birth deformities is done in five countries as shown in table 6.

**Table 6:** Newborn care during the first hour of life.

| Country* | Resuscitation<br>needed or<br>performed | Skin to<br>skin | Cord<br>care | Eye care (e.g<br>Tetracycline) | Immunization<br>(e.g. BCG) | ART/<br>NVP | Vit. K<br>inj. | Breastfed<br>within an<br>hour |
| --- | --- | --- | --- | --- | --- | --- | --- | --- |
| Argentina | X |  |  |  |  |  |  |  |
| Ghana | X |  | X | X |  |  |  |  |
| Guyana | X |  |  |  | X |  |  | X |
| Kenya |  | X | X | X | X | X | X | X |
| Malawi | X |  |  |  | X |  |  | X |
| Nigeria | X |  | X |  |  |  |  | X |
| South<br>Sudan |  |  |  | X | X |  |  | X |
| Sweden | X |  |  |  |  |  | X |  |
| Tanzania | X |  |  |  |  | X |  | X |
| Uganda | X | X | X |  | X | X | X | X |
| Zambia | X |  |  |  |  |  |  | X |

\* The seven countries whose childbirth registers were analyzed that are not shown in table 6 because the registers do not capture data on any of the eight data elements related to newborn care during the first hour of birth, are: Afghanistan, Bolivia, Eswatini, Guyana, Indonesia, Nepal, and Norway.

#### Services provided to mother prior to discharge

Health promotion intervention provided to the mother prior to discharge include counseling on infant and young child feeding in Afghanistan, Nigeria, Norway, and Tanzania. Regarding family planning counseling, only three countries capture these data (Bolivia, Nigeria, and Tanzania). Distinction is made in Zambia between exclusive breastfeeding (EBF), replacement feeding (RF), and mixed feeding (MF). In Tanzania, the type of postpartum family planning method selected by the woman is documented. In some registers, the level of detail includes whether the method is provided pre-discharge.

### Mortality and Newborn Outcomes

Childbirth or labour and delivery registers are the sources of data on maternal and newborn outcomes. Results from the analysis of maternal and neonatal death as presented for the same 18 countries.

#### Maternal death

In 14 out of 18 countries (78%), data on the status of mother at discharge (alive or died) are captured in separate columns, except for Nigeria, Sweden, and Uganda, where “maternal death” column is not included as a separate column. Data of mothers who dies are captured through an open text field. About In about half of the countries, there is open-ended space to record the cause of death, the last column in the register. In some other countries like in Tanzania, maternal deaths are recorded under “discharge” summary, the status of the mother being designated as either alive or dead.

#### Fetal and newborn death

Where the death of the baby occurs before birth, this is recorded as fetal death. Nigeria and Norway have codes to record abortion. Regarding stillbirths, 11/18 (61%) of the countries distinguish between fresh and macerated stillbirths. Bolivia and Malawi do not make such distinctions, although stillbirths are reported (Table 7). The timing of newborn death is recorded in different ways across the registers. Half the countries document early death occurring less than 7 days after delivery. In Norway which uses an electronic register, there are separate codes for recording early neonatal death (less than7 days) versus death occurring after 7 days, but before 28 days. South Sudan and Zambia have spaces to record newborn deaths whose timing is unspecified (Table 7).

**Table 7:** Abortion, fetal and early death of the newborn.

| Country | Abortion | Stillbirth<br>Fresh &<br>Macerated<br>(Combined) | Stillbirth-Fresh | Stillbirth-Macerated | Neonatal<br>death<br>(unspecified) | Early<br>death<br>(<7 days) | Death<br><28<br>days | Cause<br>of<br>death |
| --- | --- | --- | --- | --- | --- | --- | --- | --- |
| Afghanistan |  |  | X | X |  | X |  | X |
| Argentina |  |  |  |  |  |  |  |  |
| Bolivia |  | X |  |  |  |  |  |  |
| Burkina<br>Faso |  |  |  |  |  | X |  |  |
| Eswatini |  |  | X | X |  | X |  |  |
| Ghana |  |  |  | X |  | X |  |  |
| Guyana |  |  | X |  |  |  |  |  |
| Indonesia |  |  |  |  |  |  |  |  |
| Kenya |  |  | X | X |  | X |  |  |
| Malawi |  | X |  |  |  | X |  |  |
| Nepal |  |  | X | X |  |  |  |  |
| Nigeria | X |  | X | X |  | X |  |  |
| Norway | X |  | X | X |  | X | X | X |
| South<br>Sudan |  |  | X | X | X |  |  |  |
| Sweden |  |  | X | X |  | X |  |  |
| Tanzania |  |  | X | X |  |  |  | X |
| Uganda |  |  |  |  |  |  |  |  |
| Zambia |  |  | X | X | X |  |  | X |

### Postnatal Care

#### Utilization and timing of postnatal care visits post-discharge

Some countries record the date of the visit and the date of birth therefore making it possible to calculate the visit timing. In countries using cross-sectional registers, service providers count the number of visits from a mother-held record, the number of the visits a woman has attended, then enter the visit number in the postnatal care register. The timing of the PNC visit was categorized as day of birth or immediate care after birth, within 48 hours or day 2, day3-6, days 7-14, and at six weeks (Table 16). The schedule of reporting varied widely across countries, with countries like Zambia using incredibly detailed schedule as follows: 1^st^ hour, 6^th^ hour, 12^th^ hour, 18^th^ hour, 24^th^ hour, 48^th^ hour, 6^th^ day, and 6^th^ week. In some countries, the timing of the visit was unspecified.

#### Routine checks during postnatal period

Routine clinical checks, counseling, laboratory tests are done on the mother while the child receives immunization and routine checks. Blood pressure is recorded in PNC registers of five of the countries reviewed, while complications related to childbirth are recorded in seven countries. Some of the complications documented are postpartum haemorrhage, lochia (excessive or foul smelling), and presence of a fistula. Examination of the site of cesarean section is recorded in Zambia and Kenya). The mother’s hemoglobin level is recorded as such or anaemia/ pallor. Some of the less commonly documented practices include examination of the vagina, breast, and the uterus.

#### Screening and counseling services

The most common type of counseling services during postnatal visits is on family planning and nutrition counseling for both the mother and the baby. Nutrition counseling includes information on EBF, RF, and MF. Micronutrient supplementation provided mainly iron and foliate is recorded in 9/13 (70%) of the countries whose PNC registers were analyzed. Postpartum family planning is recorded in 8/13 (62%) of the countries. Screening for TB, cervical cancer, and STIs is recorded in 3, 2, and 1, countries, respectively.

#### PMTCT during the postnatal period

The postnatal period provides another opportunity for PMTCT so that infants born to HIV-infected mother remain uninfected. HIV testing is recorded in the PNC registers of six of the countries analyzed while information on whether the mother is on ART is recorded in five countries. Couple testing and the HIV status of the mother’s partner is recorded in Kenya and Zambia only (Table 8).

**Table 8:** HIV Testing and treatment services.

| Country | HIV status | HIV testing | Couple HTC | Partner HIV status | HIV results on arrival | HIV results (<6wks)/ or at discharge | On ART | On CTX |
| --- | --- | --- | --- | --- | --- | --- | --- | --- |
| Afghanistan | X | X |  |  |  |  | X | X |
| Burkina Faso | X | X |  |  |  |  |  |  |
| Eswatini | X | X |  |  | X | X | X | X |
| Kenya | X | X | X | X | X | X | X |  |
| South Sudan | X | X |  |  | X |  | X | X |
| Tanzania | X | X |  |  |  |  |  |  |
| Zambia |  |  | X | X |  |  | X |  |

#### Postnatal care for the newborn

In addition to documenting the immunizations provided to the newborn so that she or he is on schedule as per the national immunization schedule, routine checks primary focus on newborn danger signs or complications such as jaundice and failure to gain weight as expected. PNC registers of five of the countries included in the analysis record newborn danger signs.

Regarding preventing HIV infection, ART prophylaxis for the newborn is recorded in four countries (Afghanistan, Eswatini, Tanzania, and Zambia), and it is only in Eswatini where provision of early infant male circumcision is recorded.

#### Sub-analysis of registers from Ghana, Kenya, Nigeria, and Zambia

Results of analysis of registers from four countries (Ghana, Kenya, Nigeria, and Zambia) where the 2017/2018 and 2019/2020 versions of the registers were available showed that there is an apparent change towards inclusion of data elements that are critical in the computation of key maternal and newborn care indicators. Overall, 21/27(78%) key data elements for tracking maternal and newborn care are present in the latest versions of the registers reviewed (Table 9). The ones that are present in to varying degrees are: ANC visits <12 weeks, fetal heart rate, patograph use, newborn resuscitation, breast/ Cervical cancer screening and contraception counseling.

**Table 9:** Comparison of 2018 to 2020 register review form four countries.

| Category | # | Indicator / Data element | Ghana |  | Kenya |  | Nigeria |  | Zambia |  | Present in all 4 countries (2020) |
| --- | --- | --- | --- | --- | --- | --- | --- | --- | --- | --- | --- |
|  |  |  | 2018 | 2020 | 2018 | 2020 | 2018 | 2020 | 2018 | 2020 |  |
| Antenatal care | 1 | Antenatal client first visit before 12 weeks gestation |  | ✓ |  | ✓ |  |  |  | ✓ |  |
|  | 2 | Antenatal care 4th visit | ✓ | ✓ | ✓ | ✓ | ✓ | ✓ | ✓ | ✓ | ✓ |
|  | 3 | Antenatal care 8th visit |  | ✓ |  | ✓ |  | ✓ |  | ✓ | ✓ |
|  | 4 | Syphilis screening | ✓ | ✓ | ✓ | ✓ | ✓ | ✓ | ✓ | ✓ | ✓ |
|  | 5 | Syphilis test result / Treatment |  | ✓ | ✓ | ✓ | ✓ | ✓ | ✓ | ✓ | ✓ |
|  | 6 | Haemoglobin measured | ✓ | ✓ | ✓ | ✓ | ✓ | ✓ | ✓ | ✓ | ✓ |
|  | 7 | Blood pressure measurement | ✓ | ✓ | ✓ | ✓ | ✓ | ✓ | ✓ | ✓ | ✓ |
|  | 8 | Fetal heart rate | ✓ | ✓ |  |  |  |  |  |  |  |
|  | 9 | Tetanus vaccination | ✓ | ✓ | ✓ | ✓ | ✓ | ✓ | ✓ | ✓ | ✓ |
|  | 10 | PMTCT - HIV testing | ✓ | ✓ | ✓ | ✓ | ✓ | ✓ | ✓ | ✓ | ✓ |
|  | 11 | IPTp-1 for malaria during pregnancy | ✓ | ✓ | ✓ | ✓ | ✓ | ✓ | ✓ | ✓ | ✓ |
|  | 12 | IPTp-2 for malaria during pregnancy | ✓ | ✓ | ✓ | ✓ | ✓ | ✓ | ✓ | ✓ | ✓ |
|  | 13 | IPTp-3+ for malaria during pregnancy | ✓ | ✓ |  | ✓ |  | ✓ | ✓ | ✓ | ✓ |
|  | 14 | Iron supplementation / Nutrition | ✓ | ✓ | ✓ | ✓ | ✓ | ✓ | ✓ | ✓ | ✓ |
| Childbirth | 15 | Partograph used to monitor labour |  |  |  |  |  | ✓ |  | ✓ |  |
|  | 16 | Uterotonic for prevention of post-partum haemorrhage | ✓ | ✓ | ✓ | ✓ | ✓ | ✓ | ✓ | ✓ | ✓ |
|  | 17 | Newborn resuscitation / Asphyxia |  | ✓ |  |  | ✓ | ✓ | ✓ | ✓ |  |
|  | 18 | Mode of delivery/ Caesarean section | ✓ | ✓ | ✓ | ✓ | ✓ | ✓ | ✓ | ✓ | ✓ |
|  | 19 | Birthweight/ Low birth birthweight | ✓ | ✓ | ✓ | ✓ | ✓ | ✓ |  | ✓ | ✓ |
|  | 20 | Newborns breastfed within one hour of birth |  | ✓ | ✓ | ✓ |  | ✓ | ✓ | ✓ | ✓ |
| Postnatal care | 21 | Post-partum hemorrhage check/ management |  | ✓ | ✓ | ✓ | ✓ | ✓ | ✓ | ✓ | ✓ |
|  | 22 | Breast /Cervical Cancer Screening |  |  |  | ✓ |  |  | ✓ | ✓ |  |
|  | 23 | PMTCT - HIV testing | ✓ |  | ✓ | ✓ | ✓ | ✓ | ✓ | ✓ | ✓ |
|  | 24 | Contraception /FP counseling or issued | ✓ | ✓ | ✓ | ✓ |  | ✓ | ✓ | ✓ |  |
| Maternal Outcomes | 25 | Stillbirth | ✓ | ✓ | ✓ | ✓ | ✓ | ✓ | ✓ | ✓ | ✓ |
|  | 26 | Maternal death or other outcome | ✓ | ✓ | ✓ | ✓ | ✓ | ✓ | ✓ | ✓ | ✓ |
|  | 27 | Neonatal death or other outcome | ✓ | ✓ | ✓ | ✓ | ✓ | ✓ | ✓ | ✓ | ✓ |

## Discussion

This analysis reviews the registers for ANC, childbirth, and PNC for 21 countries in six regions conducted through abstracting data elements from ANC, childbirth, and PNC registers with a focus on evidence-based interventions as well as outcomes for the woman and newborn. Overall, there is great variation in the number of data elements ranging from 24 in Nepal’s childbirth register to 188 in Eswatini’s ANC register, as well as the types of elements that were captured. Countries forms also varied in terms of longitudinal versus cross-sectional data collection depending on the country context. This review has demonstrated lack of harmonization and consistency in the types of information that are collected, how they are collected and organized or recorded. Given that these registers the main source of maternal and newborn data, information gathered plays a key role in programme planning and tracking progress towards achieving national and global goals and targets.

Majority of the ANC registers reviewed for 20 countries capture routine information for prenatal care, such as gestation age, weight, height, blood pressure, and haemoglobin level. However, there were either gaps in capturing key data as per the 2016 WHO recommendations^29^ or lack of consistency in the data capture, for example on number of contacts or clinic visits, dietary interventions, and fetal assessment. Overall, few countries recorded eight or more ANC visits or having first ANC visit before 12 weeks gestation. Most of those recording eight or more ANC visits are the ones who have electronic registries, suggesting that use of electronic medical records provides an opportunity for a better tracking of ANC services. It’s noteworthy that the majority of registers (65%) included relevant medical history for the pregnant woman, although only one third included complications from previous pregnancies. Most countries captured recommended screening practices such as blood pressure measurement, hemoglobin levels and syphilis and HIV screening and treatment. However, only one third of registers captured whether an ultrasound was performed. In addition, the registers capture screening but not if any action was taken to address risk factors. For childbirth, the majority of countries capture data on obstetric complications, but few do preventative or management interventions for postpartum hemorrhage or pre-eclampsia. Although there is evidence of documentation of key data on childbirth, it is possible that some other supplemental documents such as the partograph, which is a recommended by WHO for capturing data during active labour to inform care^29^.

Regarding childbirth and PNC period, few countries tracked whether the partograph was used or fetal heart monitoring. Furthermore, data on breastfeeding within one hour of birth, newborn resuscitation and BCG immunization were also not routinely captured in most countries. The PNC registers had the fewest data elements available and the least standardization in terms of what was captured for the woman or newborn. In general, there was more information captured for the woman than for the newborn, especially around nutrition and family planning, with information for newborns focused on immunization and routine checks.

In a sub-analysis of four African countries, there was some improvement over time in terms of the types of data elements collected. These countries track the same data elements in ANC registries because of epidemiological reasons (e.g. HIV testing, STIs screening or Malaria treatment), however a great variation of information has been found in childbirth and PNC registers from these countries. Several of the newer data elements included newborn care indicators as recommended by the Every Newborn Action Plan.

There were limitations to the study. First, although every effort was made to obtain the most recent registers, these documents were not always available for all selected countries. In addition, many of the data elements reviewed were relevant to only a sub-set of countries (e.g. malaria prevention and HIV prevention and management). It was often difficult to interpret how the registers were supposed to be completed and it was not always possible to obtain instructions for completing the them, if at all they existed. Finally, we were only able to conduct a sub-analysis of four countries to assess change over time, due to limitations in obtaining the relevant registers from more countries. It would be helpful to expand this analysis to additional countries and to include the data summary forms.

Although there are global recommendations for maternal and newborn indicators to be collected within routine data systems^29–31^, this analysis highlights the great variation among and between countries. There is a need for additional guidance, standardization, and reduction of data elements to capture the most important evidence-based practices for improving the quality of care as well as tracking progress toward national and global targets. This analysis did not capture the burden on health care workers to collect these data elements in addition to providing quality care, but it is an essential consideration when selecting and updating registers and forms for routine data systems.

Improving the quality and consistency of data reported through a national HIS, requires a thorough understanding of facility registers with respect to how data recorded in facilities can be most easily translated into aggregate indicators. Taking these factors into consideration when designing a register allows for more accurate reporting and for inclusion of indicators that provide information on quality of care, such as number of service contacts and timing of interventions received.

## Conclusion

The ANC, childbirth and PNC registers are a major source of essential maternal and newborn data, but standardization in recording of data elements varies across countries. Managers and frontline health workers use these data to monitor the performance of their national health system and to guide the continuous improvement of health care services for women and newborns. The results of this analysis will complement additional work and help to inform the standardization of global indicators and provide information for other countries seeking to introduce indicators in their health systems.

## Data Availability

Summaries of registers analyzed available on Figshare: DOI:10.6084/m9.figshare.20052122

## Acknowledgement

This work was commissioned by the WHO and supported by the Bill & Melinda Gates Foundation. Special thanks to the Ministry of health officials and Jhpiego staff working in the 21 countries whose registers were analyzed for facilitating the availability of registers.

